# Multi-Omics Validation of Myelin Water Fraction as a Myelin-Specific MRI Biomarker

**DOI:** 10.1101/2025.08.27.25334582

**Authors:** Jonghyun Bae, Cassandra M. Joynes, Zhaoyuan Gong, Michael R. Duggan, Keenan A. Walker, Mustapha Bouhrara

## Abstract

The myelin sheath is critical for efficient neural signaling and central nervous system function, yet noninvasive methods to quantify myelin content *in vivo* remain limited and require robust biological validation. Myelin Water Fraction (MWF) derived from the Bayesian Monte Carlo analysis of multicomponent Driven Equilibrium Single Pulse Observation of T_1_ and T_2_ (BMC-mcDESPOT) imaging method offers a promising biomarker of myelin content, but its biological specificity requires further validation. To elucidate the molecular underpinnings of MWF derived using BMC-mcDESPOT, we integrated this advanced MRI method with high-throughput plasma proteomics and spatial transcriptomics across major white and deep gray matter regions in adults. Plasma protein analyses using the SomaScan 7k platform revealed significant associations between MWF and proteins highly expressed by oligodendrocytes, a key cell type for myelin synthesis and maintenance. Furthermore, spatial mapping of Myelin Basic Protein (*MBP*) gene expression obtained from the Allen Human Brain Atlas demonstrated a strong positive correlation with regional MWF values, consistent with known myelin distribution patterns. These convergent findings validate MWF as a biologically relevant, myelin-specific imaging marker and highlight the potential of integrating omics data to validate and enhance *in vivo* neuroimaging biomarkers. Our work evinces and advances the utility of MWF for studying white matter integrity in health and disease, offering new avenues for clinical and translational neuroscience.

## Introduction

The myelin sheath is essential for efficient neural signaling and central nervous system (CNS) function. It facilitates saltatory conduction and supports axonal integrity, playing a crucial role in both development and adult brain health. Disruption or loss of myelin is a hallmark of numerous neurological disorders, including multiple sclerosis, leukodystrophies, and neurodegenerative diseases such as Alzheimer’s disease [1–3]. Given its fundamental role, the ability to non-invasively assess myelin content *in vivo* is a high priority in both clinical and research settings.

One established approach for imaging myelin is through the quantification of myelin water fraction (MWF), which reflects the proportion of water trapped between the layers of the myelin sheath. Among the various MRI methods developed to estimate MWF, multicomponent Driven Equilibrium Single Pulse Observation of T_1_ and T_2_ (mcDESPOT) has gained traction due to its whole-brain coverage and relatively short acquisition times [4–6]. More recently, Bayesian Monte Carlo analysis of mcDESPOT (BMC-mcDESPOT) has greatly improved MWF estimation by incorporating prior distributions and modeling uncertainty [7]. While MWF obtained via BMC-mcDESPOT techniques shows regional patterns consistent with known myeloarchitecture, its cellular and molecular specificity for myelin content remains under investigation.

Historically, validation efforts have relied on histological comparisons in post-mortem tissue or on indirect associations with demyelinating pathology. However, histology, often regarded as the “gold standard”, is inherently destructive, with tissue undergoing significant denaturation during processing. Further, formalin fixation has been shown to drastically alter MWF values, introducing substantial bias [8, 9]. These limitations highlight the need for more innovative, non-destructive methods to validate the specificity of MRI-derived myelin biomarkers.

To address this gap, we conducted an extensive validation of BMC-mcDESPOT derived MWF using omics-based approaches. Specifically, we integrated proteomic and transcriptomic analyses with neuroimaging data to examine the molecular correlates of MWF across the brain. Leveraging data from a cohort of 85 participants, we used the SomaScan platform to measure 6,393 plasma proteins and assessed their relationships with regional MWF. Results revealed significant associations between MWF and proteins involved in white matter synthesis as well as the function and metabolism of oligodendrocytes, supporting the biological relevance of our imaging measure. Additionally, we demonstrated strong regional correlations between aggregated MWF levels, derived from a cohort of 137 participants, and mRNA levels of a canonical myelin-related gene, myelin basic protein (*MBP*), derived from data aggregated over 6 donors [10]. Together, these findings offer a novel molecular- and neurobiology-informed validation of MWF derived from our BMC-mcDESPOT method. By integrating high-throughput omics data with quantitative neuroimaging, this work supports the hypothesis that MWF reflects underlying myelin biology and thereby represents an essential step toward establishing MWF as a robust and specific biomarker of myelin *in vivo*.

## Methods

### Myelin Water Fraction MRI

This study included 137 subjects from the Baltimore Longitudinal Study of Aging (BLSA) (age = 22-94, M/F=77/60). To quantify MWF, each subject underwent the BMC-mcDESPOT protocol, consisting of 3D spoiled gradient echo (SPGR) sequences (TE/TR = 1.48/5 ms) and balanced steady state free precession (bSSFP) sequences (TE/TR = 2.8/5.9 ms) with RF excitation pulse phase increments of 0 and *π* to account for off-resonance artifacts [4]. Imaging was performed with an acquisition matrix of 150 × 130 × 94 and an isotropic voxel size of 1.6 mm, then reconstructed to 1 mm isotropic resolution. To correct for RF inhomogeneity (*B*_1_), the double-angle method [11] was applied, using two fast spin-echo images with flip-angles of 45° and 90° (TE/TR = 102/3000 ms, spatial resolution = 2.6 × 2.6 × 4 *mm*). The estimated low-resolution *B*_1_ field was then interpolated to match the spatial resolution of the SPGR and bSSFP images. MWF was estimated based on a two-compartment, non-exchanging model that distinguishes between short and long T_l_ and T_2_ components. The short component reflects water confined within the myelin sheaths, whereas the long component represents intra- and extracellular water. We identified MWF values in major white matter tracts, such as the corpus callosum, internal capsule, and corona radiata, using the JHU white matter atlas [12]. We also obtained MWF values in deep gray matter structures, such as the caudate nucleus, putamen, and thalamus, using the Desikan-Killiany atlas [13].

### Plasma Proteomics

We investigated the relationship between 6,393 plasma proteins, measured using the SomaScan v.4.1 proteomic platform (SomaLogic), and whole-brain white matter MWF measurements. SomaScan quality control procedures for the BLSA study have been described previously [14, 15]. Linear regressions examined the association of log2 transformed protein levels with MWF after adjusting for age at protein measurement, sex, estimated glomerular filtration rate (eGFR) [16], and age^2^. The age^2^ term was included to account for nonlinear patterns of change in MWF with age, as previously reported [17]. The Human Protein Atlas (HPA) [18], a publicly available dataset, was used to annotate the cell-specificity of candidate proteins [19]. This analysis included 85 participants with both plasma proteomics and MWF measures (Fig. 1).

**Figure 1.**
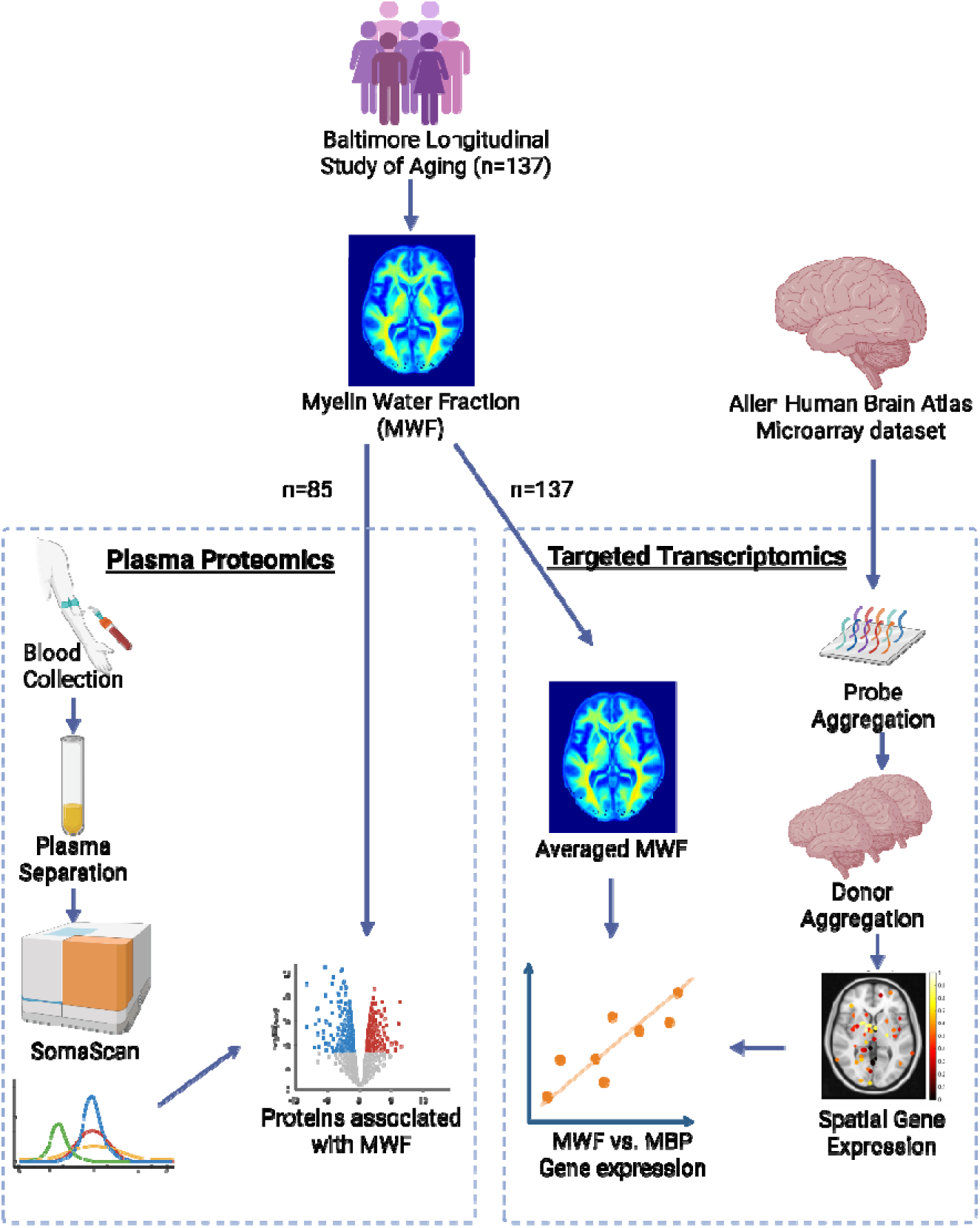
Study design for proteomics and transcriptomics. For the proteomic analysis, we selected a subset of participants from the Baltimore Longitudinal Study of Aging (BLSA) who had both brain imaging and blood samples available. Plasma protein levels were measured using the SomaScan 7k platform. Proteins significantly associated with whole-brain white matter Myelin Water Fraction (MWF) were identified, and cell-type specificity was annotated using expression data from the Human Protein Atlas (HPA). For the transcriptomic analysis, we focused on Myelin Basic Protein (MBP) mRNA levels and its spatial correlation with regional MWF measurements. MBP mRNA levels were obtained from the Allen Human Brain Atlas (AHBA) microarray dataset.

Plasma protein-based cell-specificity annotations were derived following the approach described in a previous study [20]. Normalized transcripts per million (nTPM) were obtained from the HPA for the gene corresponding to each protein. For each gene, nTPM were additionally normalized across all available cell types via z-scoring using the geometric mean and standard deviation. This step accounts for non-linear expression distributions and enables standardized comparisons of cell-specificity across proteins, which may have differential levels of baseline expression. Proteins with genes whose z-scores were ≥ 2 in brain-related cell types were considered to have enriched expression by that cell type. Proteins enriched in more than one brain-related cell types were designated as oligoMultiCell when oligodendrocytes or oligodendrocytes precursor cells (OPCs) were included among the enriched types, and as multiCell when they are not.

Following linear regression, enrichment analysis was performed separately for all proteins that exhibited significant (p < 0.05) positive (n=106) or negative (n=101) associations with MWF. In essence, enrichment analysis takes a given gene set list and finds canonical biological pathways with a high degree of overlap. For our enrichment analysis we used the website Enrichr (https://maayanlab.cloud/Enrichr/) [21–23] and focused on results from seven annotation databases, for each database we report the top 5 most significantly enriched pathways or all pathways with a p < 0.05, whichever is fewer.

### Transcriptomics

To map the spatial distribution of the Myelin Basic Protein (*MBP*) gene, we utilized the Allen Human Brain Atlas (AHBA) microarray dataset, which measures genome-wide messenger RNA (mRNA) levels from a total of 3,700 brain tissue samples across six adult donors, encompassing cortical, subcortical, brainstem, and white matter regions [24]. Data processing was conducted using the abagen toolbox [25]. Representative probe selection from the four available probes corresponding to the *MBP* gene was based on differential stability [10], calculated as the average Spearman correlation of each probe’s expression across brain regions between all donor pairs. The probe with the highest mean correlation was retained. Finally, mRNA levels were aggregated among donors, and the resulting gene expression profiles were mapped onto the same atlases used to delineate regional MWF values (i.e. JHU white matter and Desikan-Killiany atlas) based on their MNI coordinates. When assessing the regional *MBP* mRNA levels, samples were assigned to an ROI when placed within 2 mm of each parcellation. We obtained *MBP* levels across major white matter tracts and deep gray matter structures, identical to those obtained from MWF maps. We correlated our regional mean MWF measurements, derived from the aggregated maps across all 137 participants, with the aggregated regional mRNA levels for each brain region using a linear regression with mean regional MWF as the dependent variable and mean regional mRNA levels as the independent variable (Fig. 1).

## Results

Table 1 summarizes the demographic characteristics of the dataset used in this study. Our proteomics study utilized a subset of BLSA participants (n=85) who completed both MR imaging and blood collection protocols. For the transcriptomics analyses, we included all BLSA participants who underwent MR imaging (n=137). The participants were well-balanced in terms of sex distribution (56% male) and there were no significant differences in age between males and females (*p* > 0.1). Racial distribution was predominantly White (70 %), followed by Black (19%). The AHBA microarray dataset comprises mRNA levels from six adult human donor brains, predominantly male, aged 24 - 57 years.

**Table 1.** Participant Demographics.

|  | Proteomics | Transcriptomics |  |
| --- | --- | --- | --- |
|  | Baltimore Longitudinal Study of Aging (BLSA) | BLSA | Allen Human Brain Atlas Microarray dataset |
| Characteristics | <u>Cognitively normal</u> (CN) participants (n = 70)<br><u>Mild Cognitive Impairment</u> (n = 6)<br><u>Dementia</u> (n = 6)<br><u>Non-Dementia Cognitive Impairment</u> (n = 3)<br>Total = 85 | CN participants (n = 137) | 6 healthy adult donors |
| Age, mean (SD) | At Protein Measurement 58.0 (21.68)<br>At MWF Measurement 60.2 (22.5) | 59.1 (21.3) | 42.5 (13.4) |
| Sex, Men (%) | 45 (53.0%) | 77 (56.2%) | 5 (83.3%) |
| Race, White (%) | 55 (64.7%) | 96 (70.1%) | 3 (50.0%) |
| Race, Black (%) | 19 (22.4%) | 26 (19.0%) | 2 (33.3%) |

Figure 2 presents a volcano plot that illustrates the relationship between each measured plasma protein and global white matter MWF values. When applicable, each protein is annotated with the CNS cell type(s) where that protein is highly expressed. The plot reveals a spectrum of protein-MWF associations, highlighting 106 proteins that are significantly more (unadjusted p < 0.05) abundant and 101 proteins that are significantly less (unadjusted p < 0.05) abundant in the plasma of participants with greater whole-brain white matter MWF. Proteins such as ADAM10, ADGRB1, and RYBP were positively associated with MWF and are implicated in white matter development and maintenance, particularly the function and metabolism of oligodendrocytes and OPCs – the primary glial cells involved in myelination. Since oligodendrocytes are responsible for the formation of the myelin sheath, the primarily positive associations between MWF and proteins highly differentially expressed by oligodendrocytes are biologically consistent. Full details of the regression results for all proteins are provided in the Supplementary material.

**Figure 2.**
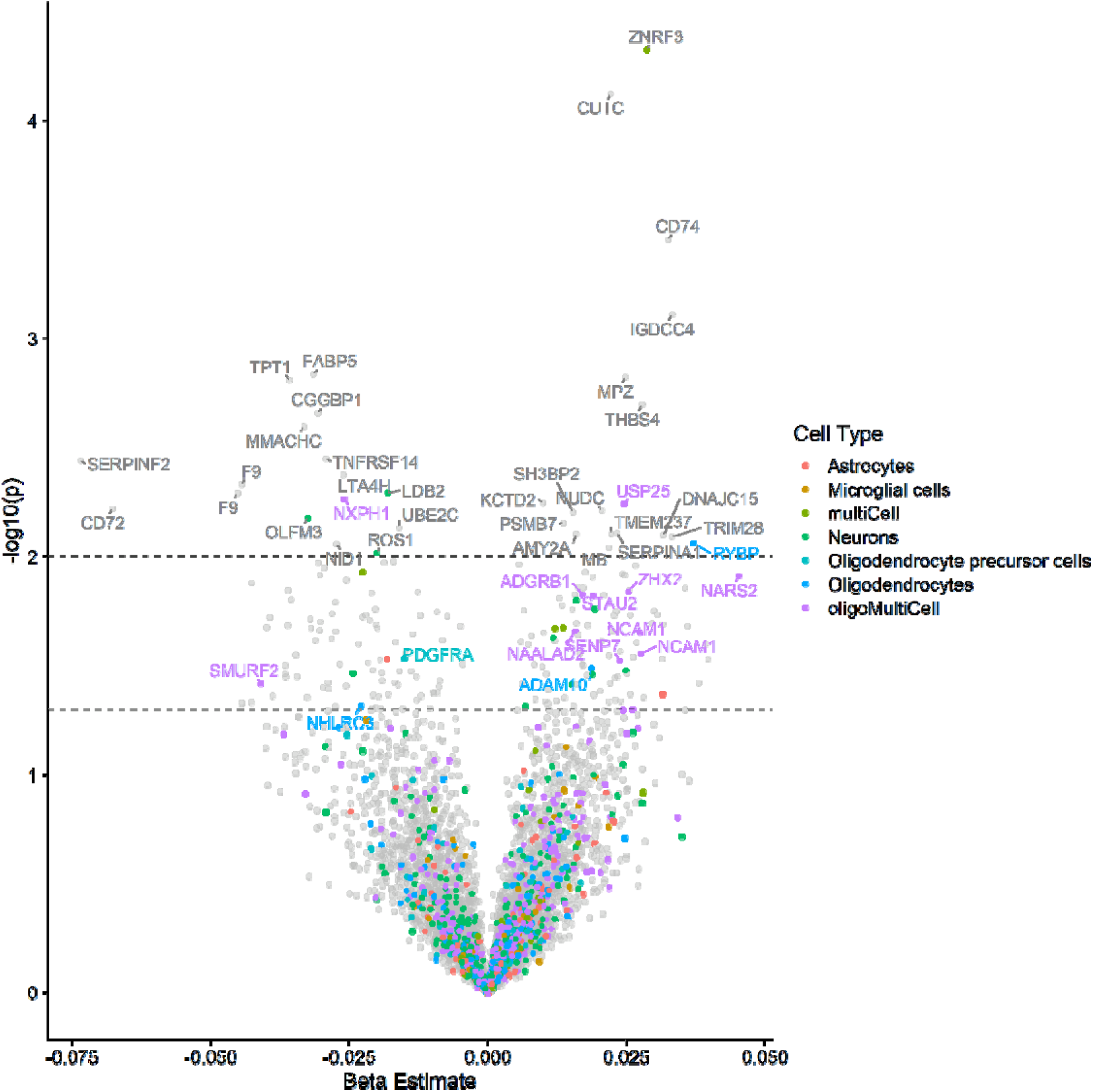
Volcano plot displaying the correlation between plasma proteins and MWF measurements. The x axis indicates the magnitude and direction of the association between the protein abundance and MWF derived from adjusted linear regression models; the y axis is the - log10 of the p-value for that β term in the regression, higher values indicate a more significant relationship. When applicable, proteins are annotated with the cell type(s) by which it is differentially expressed. 15 of the proteins with nominally significant p-values () are highly expressed by oligodendrocytes (n=3), oligodendrocyte precursor cells (n=1), or by at least two cell types, of which at least one was either oligodendrocytes or oligodendrocyte precursor cells (n = 11). Dashed lines indicate nominal significance levels of p<0.05 and p<0.01.

The majority of the MWF-associated proteins were not brain cell specific (175 out of 207), but of the 32 proteins with cell specific expression patterns, 15 were highly expressed in either (or both) oligodendrocytes or OPCs. To better understand the biological processes related to MWF, we conducted enrichment analyses of all significant MWF-related proteins using complementary and publicly available databases. The top pathways consisted of several immune and digestion/metabolic pathways (Figure 3). Of note, adaptive immune pathways, B and T cell activation and MHC class II presentation, are enriched among proteins positively (right hand side) associated with MWF, whereas inflammation, complement signaling, and other aspects of the innate immune response are enriched among proteins negatively associated with MWF. In sum, the enrichment results suggest that adaptive immune function and adequate regulation of bioenergetic processes may be among the peripheral processes most relevant to preserving myelin content.

**Figure 3.**
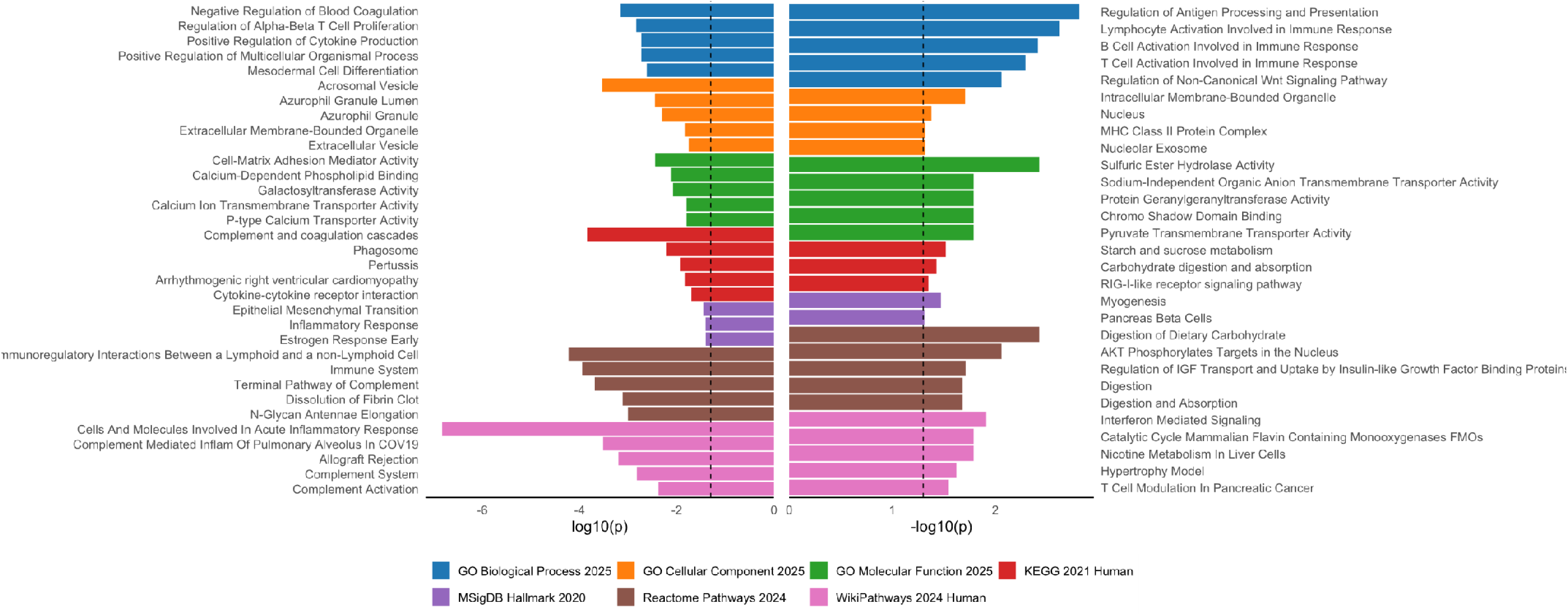
Pathways identified via enrichment analysis of proteins significantly (p < 0.05) positively (n=106, right-hand side) or negatively (n=101, left-hand side) associated with MWF. Bars indicate the log10 transformation of the p-value; larger bars indicate a greater significance in the overlap between the MWF associated protein list and the canonical pathway listed. For each annotation database the top 5 pathways or all pathways with p < 0.05 are listed, whichever is fewer. Dashed line indicates p = 0.05.

Figure 4 displays a spatial correlation plot comparing regional *MBP* mRNA levels with corresponding MWF values across major white matter tracts and deep gray matter regions. MWF values are substantially lower in deep gray matter regions, represented by darker-colored markers, as these areas primarily consist of neural cell bodies and thus are less myelinated. In contrast, major white matter tracts, indicated by lighter-colored markers, exhibit higher MWF values consistent with their dense myelin content. This spatial pattern is paralleled in the gene expression data: *MBP* levels are markedly reduced in gray matter regions and elevated in white matter tracts. We find a robust positive correlation (r=0.69; *p*=0.012) between *MBP* mRNA levels and MWF values across brain regions, reinforcing the role of *MBP* as a molecular marker of myelin content and validating MWF as a reliable *in vivo* imaging metric for assessing regional myelination.

**Figure 4.**
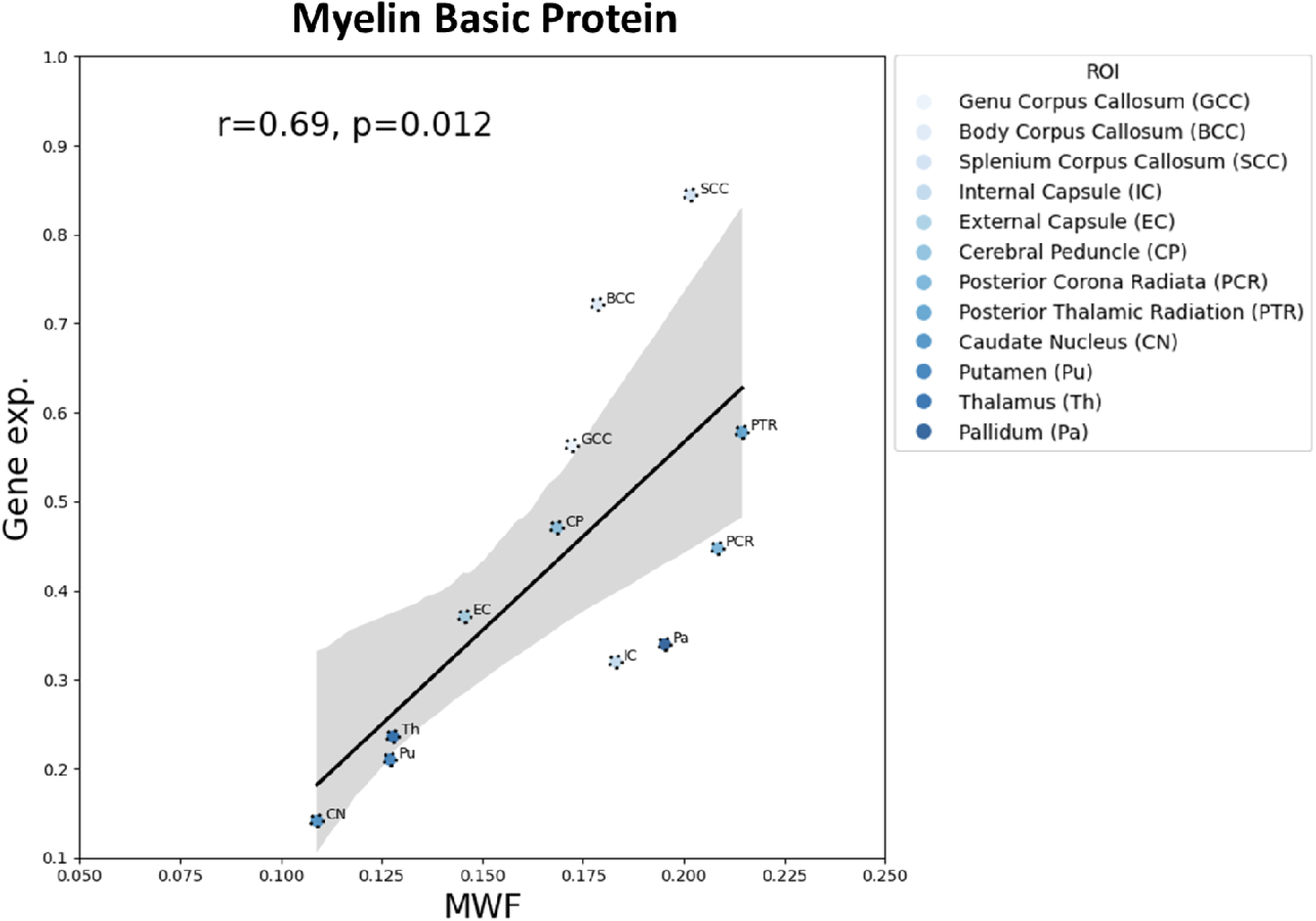
Correlation plot between Myelin Basic Protein (MBP) and Myelin Water Fraction (MWF) measurements in different brain regions. Microarray data were obtained from the Allen Human Brain Atlas, which contains the gene expression data from 6 healthy adult donors.

## Discussion

In this study, we leveraged high-throughput proteomic and transcriptomic data to validate the biological specificity of MWF derived from the BMC-mcDESPOT MRI protocol to myelin content. Our findings demonstrate that global MWF values are strongly associated with proteomic markers of myelin content, particularly those related to oligodendrocyte function. We also demonstrate a strong correlation between regional levels of MWF and *MBP* mRNA levels, assessed in region specific post-mortem brain samples across various brain regions, including deep gray matter and major white matter tracts. Together, this multimodal data provides additional support for MWF as a tool to measure facets of myelin biology *in vivo*, advancing its utility as a quantitative biomarker for white matter myelination and integrity.

Consistent with the known cellular architecture of the central nervous system, we observed that plasma protein levels most strongly correlated with MWF were highly expressed by cell types involved in myelination, particularly oligodendrocytes and OPCs [26]. These cells are critically important to the production and maintenance of the myelin sheath, and their activity is closely tied to myelin turnover and white matter plasticity throughout the lifespan. Although the majority of the proteins significantly associated with MWF (unadjusted p < 0.05) were not specific to CNS cells, of those that had cell-specific expression patterns, many (46.9%) were specifically expressed by oligodendrocytes, OPCs, or oligodendrocytes and/or OPCs in conjunction with another CNS cell type. The significant positive associations between MWF and proteins related to oligodendrocyte maturation and myelination, such as ADAM10 [27, 28], support the proposition that our imaging measure is sensitive to cellular processes involved in myelin synthesis and maintenance. In addition to the oligodendrocyte and OPC specific proteins associated with MWF, several of the significant proteins in our regression analysis were highly expressed by neurons (n=7 and n=4 neuron specific proteins positively and negatively associated with MWF, respectively), suggesting that MWF may capture broader aspects of neuronal health and intercellular interactions within the myelin microenvironment, providing potential proteomic targets for future myelination-related investigations.

Our enrichment analyses separately using the proteins significantly positively or negatively associated with MWF provide additional confirmation of the biological specificity of this measure. Proteins which were positively associated with MWF were annotated to adaptive immunity related pathways, such as “B Cell activation Involved in Immune Response” and “T Cell Activation Involved in Immune Response”, as well as to digestive and metabolic pathways, such as “Digestion” and “Carbohydrate digestion and absorption”. Additionally, proteins negatively associated with MWF were annotated to innate immunity related pathways such as “Cells and Molecules Involved In Acute Inflammatory Response” and “Inflammatory Response”. These enrichment results are consistent with previous findings which have shown a relationship between infection and white matter integrity [29], and between diet and white matter levels [30].

The spatial correspondence between MWF and *MBP* mRNA levels further strengthens the biological validity of MWF as a myelin-specific marker across brain regions. *MBP* is one of the most abundant proteins in the myelin sheath and plays a key role in stabilizing the multilamellar structure of compact myelin [31]. Using transcriptomic data from the Allen Human Brain Atlas, we found that *MBP* mRNA levels resembled the spatial distribution of MWF values, with consistently higher expression in white matter tracts and lower in deep gray matter regions. The robust positive correlation between regional *MBP* mRNA levels and MWF provides converging molecular evidence that BMC-mcDESPOT MWF is sensitive to the density and distribution of myelin at the transcriptomic level. This cross-modality consistency is critical, given the absence of reliable *in vivo* histological comparators for human myelin quantification.

Importantly, our results extend previous validation efforts by incorporating non-invasive, scalable -omics approaches that do not suffer from the limitations of traditional histology. Post-mortem validation techniques are inherently destructive and can introduce significant artifacts, particularly in the context of tissue fixation and processing [32, 33]. Our approach circumvents these issues by leveraging peripheral proteomic signatures and spatial gene expression atlases to interrogate the biological underpinnings of imaging markers in living individuals. This integrative framework sets the stage for future work combining neuroimaging with systems biology, particularly in large-scale population studies and clinical trials targeting white matter integrity.

Despite these strengths, several limitations should be acknowledged. First, the associations between plasma proteins and brain-derived MWF are necessarily indirect, given the blood-brain barrier and the complex relationship between peripheral biomarkers and CNS pathology. While the cell-specificity analysis helps contextualize the biological relevance of these proteins, causality cannot be inferred. Additionally, although the Allen Brain Atlas provides high-resolution spatial transcriptomic data, it is derived from a limited number of adult donors and does not fully capture inter-individual variability in gene expression. Finally, while BMC-mcDESPOT improves upon earlier MWF estimation methods by modeling uncertainty and incorporating priors, this technique still relies on assumptions regarding tissue compartments and relaxation properties that may not fully capture the heterogeneity of human white matter [34].

In conclusion, our study supports the utility of BMC-mcDESPOT-derived MWF as a biologically meaningful and myelin-sensitive metric. By integrating imaging, proteomic, and transcriptomic data, we demonstrate that MWF reflects not only structural features of white matter but also the underlying transcriptomic and proteomic activity associated with myelin-forming cells (namely oligodendrocytes). These results support the use of MWF as a non-invasive biomarker for assessing white matter myelination and health across a range of neurological conditions, illuminating new avenues for exploring the molecular correlates of brain microstructure *in vivo*.

## Data Availability

The datasets generated and/or analyzed during this study are available upon reasonable request to the corresponding author, contingent upon institutional approval.

